# Effects of Age and Sex on Systemic Inflammation and Cardiometabolic Function in Individuals with Type 2 Diabetes

**DOI:** 10.1101/2024.05.08.24307092

**Authors:** Jody Dushay, Eva S. Rickers, Enya Wang, Jessica Gilman, Ying Zhang, Ron Blankstein, Ernest V. Gervino, Michael Jerosch-Herold, Aristidis Veves

## Abstract

**Objective:** Systemic inflammation, aging, and type 2 diabetes (T2DM) all contribute to the development of cardiovascular dysfunction and impaired aerobic exercise capacity but their interplay remains unclear. This study evaluates the impact of age, sex, and inflammation on coronary and peripheral vascular function and exercise capacity in elderly individuals with and without type 2 diabetes (T2DM).

**Research Design and Methods:** Elderly individuals (age ≥65 years) underwent biochemical and tissue inflammatory phenotyping, cardiopulmonary exercise testing (CPET), cardiovascular magnetic resonance (CMR) imaging, and vascular reactivity testing. Correlation and regression analyses determined the effects of systemic inflammation, older age, and sex on cardiovascular health, stratified by T2DM status.

**Results:** For the 133 recruited individuals (44% female; median age 71, IQR=7 years, 41% with T2DM) the presence of T2DM did not have an effect on most blood serum inflammatory markers and skin biopsies. Hyperemic myocardial blood flow (hMBF), flow-mediated, and flow-independent nitroglycerin induced brachial artery dilation were significantly impaired in males, but not females with T2DM. Peak VO2 was lower with T2DM (p=0.022), mostly because of the effect of T2DM in females. Females showed more adverse myocardial remodeling assessed by extracellular volume (p=0.008), independent of T2DM status.

**Conclusions:** Our findings suggest that the pathophysiological manifestations of T2DM on vascular function and aerobic exercise capacity are distinct in elderly males and females and this may reflect underlying differences in vascular and myocardial aging in the presence of T2DM.

## INTRODUCTION

Cardiovascular disease (CVD) is a leading cause of death among older individuals with type 2 diabetes mellitus (T2DM) ^1,2^. Given its role as a shared pathophysiological hallmark of aging, diabetes, and CVD, systemic inflammation has been a major target of ongoing efforts to mitigate CVD risk in diabetes ^3,4^. Most effects of systemic inflammation on the vasculature manifest phenotypically at the level of the microvasculature, where increased oxidative stress, immune system activation, and epigenetic factors converge to impair microvascular function across coronary and non-coronary beds ^5,6^. These effects are amplified with aging ^7^.

The multifaceted changes in cardiovascular structure and function with aging, termed “cardiovascular aging,” reflect a complex interplay among cumulative clinical risk factors (e.g. obesity, diabetes), modifiable lifestyle traits (e.g. physical activity, diet), and inflammation ^8^. yet comprehensive imaging-based studies in older individuals at risk for advanced microcirculatory cardiovascular conditions (particularly those with diabetes) are limited. In this context, cardiovascular magnetic resonance (CMR) offers a capability to assess myocardial blood flow (an index of microcirculatory coronary function) alongside broad organ– and tissue-level myocardial phenotyping (LV mass and volumes, tissue fibrosis) relevant to cardiovascular aging. It remains uncertain whether optimal medical management of CVD risk factors in individuals with T2DM effectively preserves coronary function as compared to age-matched individuals without T2DM, and whether systemic inflammation impairs coronary function similarly in elderly individuals with and without T2DM. Addressing this issue is essential, as the impact of systemic inflammation on coronary function in these specific populations remains uncertain. Additionally, although data indicate increased risk of CVD among females after menopause ^9,10^, there are few studies that focus specifically on females who are 10 or more years post-menopausal.

The goal of this study was to investigate the effects of T2DM, advanced age and sex on vascular function, systemic inflammation, and cardiopulmonary exercise capacity. To that end, we performed extensive cardiovascular phenotyping using innovative techniques including a) biochemical assays of cytokines, chemokines and proinflammatory mediators of immune activation and inflammation in blood serum and skin biopsies ^11,12^, b) CMR imaging for assessment of ventricular volumes and function, tissue characterization by T1 mapping ^13–15^, rest and stress myocardial perfusion imaging for myocardial blood flow quantification, c) cardiopulmonary exercise testing (CPET), and d) vascular reactivity of the brachial artery and skin. CPET measures VO2 max during exercise, a measure of cardiovascular fitness, and provides information about cardiopulmonary and muscle physiology ^16^.

## METHODS

### Study Population

We prospectively recruited through local clinics and community advertisements individuals of age 65 years and older, with and without T2DM. The diagnosis of T2DM followed published ADA criteria ^17^. Individuals with contraindications to CMR, LV systolic dysfunction (defined as LV ejection fraction < 50% by any imaging modality within 1 year prior to enrollment) and prior ST-elevation myocardial infarction by clinical history or electrocardiographic Q waves (in at least 2 contiguous leads) were excluded. Individuals with HbA1c ≤6.5 % and without previous diagnosis of diabetes were enrolled as healthy volunteers. Additional exclusion criteria were current cigarette or tobacco use, unstable cardiac conditions, severe COPD, use of oral anticoagulant therapy, GFR <50 ml/min, prior hemorrhagic stroke, and any other condition possibly preventing completion of the CPET-study protocol ^18^.

The initial study visit included review of medical history, physical examination, completion of dietary and physical activity questionnaires assessing baseline self-reported metabolic equivalents (METs) ^19^, urine collection, blood draw, vascular reactivity testing, and skin biopsy. A subsequent study visit within 2 weeks included CMR, CPET. A 50 ml blood sample (∼35 ml plasma, ∼10 ml serum, and a buffy coat in a PAXgene tube) was collected at the screening and follow up visits. Blood was processed via ultracentrifugation and stored at –80 °C. The study was approved by the Institutional Review Board of the Beth Israel Deaconess Medical Center and written informed consent was obtained from all subjects.

### Stress Cardiac Magnetic Resonance Imaging (CMR)

CMR studies were performed after an overnight fast and 24-hour abstinence from caffeine intake with a 3 Tesla MRI scanner (Siemens Medical Solutions, Erlangen, Germany) with a dedicated 18-element phased body array. The CMR protocol included: a) assessment of ventricular volumes and function with steady state free precession (SSFP) cine imaging; b) first pass contrast-enhanced T1-weighted myocardial perfusion imaging in three short-axis slices (basal, mid, apical levels) through the LV; c) T1 mapping with a modified Look-Locker imaging (MOLLI) sequence before and after gadolinium contrast administration for assessment of extracellular volume expansion, a maker of diffuse fibrosis; and d) imaging of late gadolinium enhancement (LGE) to detect any myocardial scar ^13–15^. A contrast bolus dosage of 0.06 mmol of Gd-DTPA (Magnevist, Berlex Inc.) was injected approximately 5 seconds from the start of the perfusion scan, and a total of 80 images were acquired per slice. Rest perfusion imaging was performed first, followed 20 minutes later by stress perfusion imaging after inducing hyperemia by bolus injection of 0.4 mg/5-mL of regadenoson over 10 seconds (Astellas Pharma US, Inc., Northbrook, IL). After both stress and rest perfusion imaging, an additional gadolinium dose was injected to reach a total of 0.15 mmol/kg before post-contrast T1 mapping.

Cine, perfusion, MOLLI, and LGE images were analyzed with QMASS CMR image analysis software (QMASS version 9.3, Medis Medical Imaging Systems, Leiden, The Netherlands) by tracing the endocardial and epicardial contours avoiding the blood pool. Myocardial blood flow (MBF) was quantified in ml/min/g for 16 myocardial segments in three short-axis slices. Using MOLLI T1 mapping, segmental myocardial R1 (1/T1) was plotted against blood R1, to determine the extracellular volume fraction (ECV) using the (same day) measured blood hematocrit. Global longitudinal strain (GLS) of the left ventricle was quantified by feature tracking on long-axis cine images (QStrain, Medis Medical Imaging Systems, Leiden, The Netherlands).

## CPET

An IV catheter was placed in an arm vein and 20cc of blood was collected at rest. Subjects then exercised on a stationary bike using a ramp protocol (10-15 watts per stage) until peak volitional fatigue and/or respiratory exchange ratio (RER) >1.20 at which time a second sample of 20cc of blood was collected. Continuous ventilatory expired gas analysis was obtained (Medgraphics Ultima CPX Metabolic Stress Testing system, Minneapolis, Minnesota). The oxygen and carbon dioxide sensors were calibrated using gases with known oxygen and carbon dioxide concentrations before each test. The flow sensor was calibrated with a 3L syringe before each test. VO2 (mL/kg/min), VCO2 (L/min), and VE (L/min) were collected throughout the exercise test ^20^. The respiratory exchange ratio (RER) was calculated by dividing the carbon dioxide output (VCO2) by VO2. The chronotropic index (CI) for CPET was calculated as difference of peak heart rate during exercise and resting heart rate, normalized by the age-adjusted expected heart response, defined as 220 – age [years] – resting heart rate.

### Vascular function testing

Endothelial function was assessed by measuring flow mediated dilation (FMD) in accordance with published guidelines ^21^. Briefly, brachial artery diameter was measured before and during reactive hyperemia using high resolution ultrasound with a 10.0 MHz linear array transducer and an Aloka Prosound 7 system (Hitachi Aloka Medical Ltd, Tokyo, Japan). Reactive hyperemia was induced by inflating a pneumatic tourniquet distal to the brachial artery to 50 mmHg above systolic blood pressure for 5 minutes and then deflating it rapidly. FMD was expressed as the percentage change between baseline and post-occlusive artery diameter. For nitroglycerin-induced vasodilation (NID), a third and fourth ultrasound of the brachial artery were performed, after 20 minutes of supine rest and again 4-minutes after administration of 400 µg sublingual nitroglycerin (NTG). NID was expressed as the percentage change between baseline and post-nitroglycerin brachial artery diameter ^21–23^. NID was not performed in 19 subjects with T2DM and 17 without due to contraindication to NTG administration.

Skin blood flow was measured in a temperature-controlled environment before and after iontophoresis of acetylcholine (ACh; endothelium-dependent) and sodium nitroprusside (SNP; endothelium-independent) using laser Doppler flowmetry (DRT4 Laser Doppler Blood Flow Monitor, Moor Instruments Ltd, Millwey, Devon, England) ^24^. The percentage change in skin blood flow after ACh and SNP administration compared to baseline were used as measures of micro-circulatory function.

### Skin biopsy

Two 3 mm punch skin biopsies at the volar aspect of the forearm were performed for each participant. The specimen was placed directly into a Cryomold pre-filled with OCT (Tissue-Tek®) solution, stored at –80 °C and used for immunohistochemistry and immunofluorescent measurements to analyze mast cell degranulation and macrophage polarization.

### Statistical Analysis

The primary endpoints were differences in inflammatory markers, vascular reactivity, cardiac function and exercise capacity between elderly individuals with and without T2DM. Data were expressed as mean ± standard deviation for normally distributed data or median with interquartile range (IQR) for non-normally distributed data. Between-group comparisons were performed using the Student’s t-test or non-parametric tests (Mann-Whitney) when data were not normally distributed. Relationships between quantitative variables were assessed with Pearson’s correlation test or Spearman’s rank correlation as applicable. The effects of T2DM, sex and their interaction were analyzed by 2-way ANOVA, followed by *post hoc* pairwise comparisons of marginal means using Tukey’s method for adjustment of p-values. The Levene test was used to verify the homogeneity of variance across groups in the ANOVA model. Peak VO2 was analyzed with a linear regression model that included age, BMI, and sex as predictors, as with widely used models for prediction of peak VO2 ^25^. Additional predictors were T2DM status and chronotropic index (CI) to adjust for chronotropic incompetence, and the interactions of BMI and CI with sex. A similar multi-variable linear regression model was built for analyzing peak O2 pulse, but here myocardial ECV was added as predictor to determine any effects of myocardial remodeling on stroke volume. A *p*-value of <0.05 was considered statistically significant. Statistical analyses were performed using SPSS Version 23.0 (IBM Corp., Armonk, NY, USA), the R environment (R version 4.1.2, R Foundation for Statistical Computing, Vienna, Austria) and the Minitab Statistical Package Version 21.3 (Minitab Inc., State College, PA, USA).

## RESULTS

### Study population baseline characteristics

We enrolled 133 individuals, 54 with T2DM and 79 without T2DM (**Table 1**). There were no differences in age, sex, race, or ethnicity between the groups. T2DM participants had higher BMI (p=0.11) and waist to hip ratio (p=0.007), lower diastolic blood pressure (p=0.021), and lower total and LDL cholesterol (both p<0.0001). Statin and antihypertensive medication use were significantly higher among individuals with T2DM compared to controls (p<0.0001 for both). There were no differences between controls and T2DM in baseline self-reported metabolic equivalents (METs), either for vigorous or moderate exercise or all activities (Table 2). There were no differences in METs between males and females.

**Table 1.**
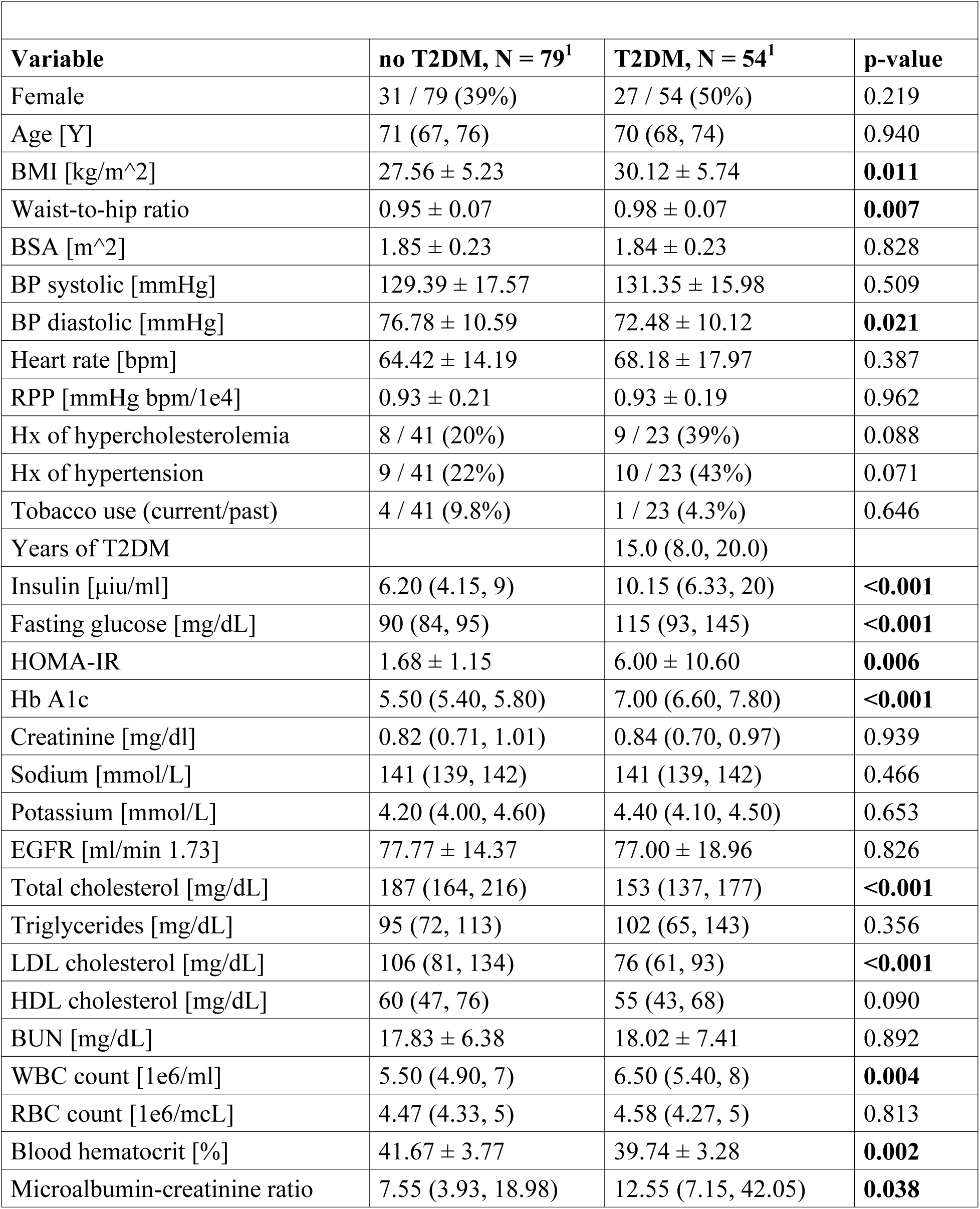
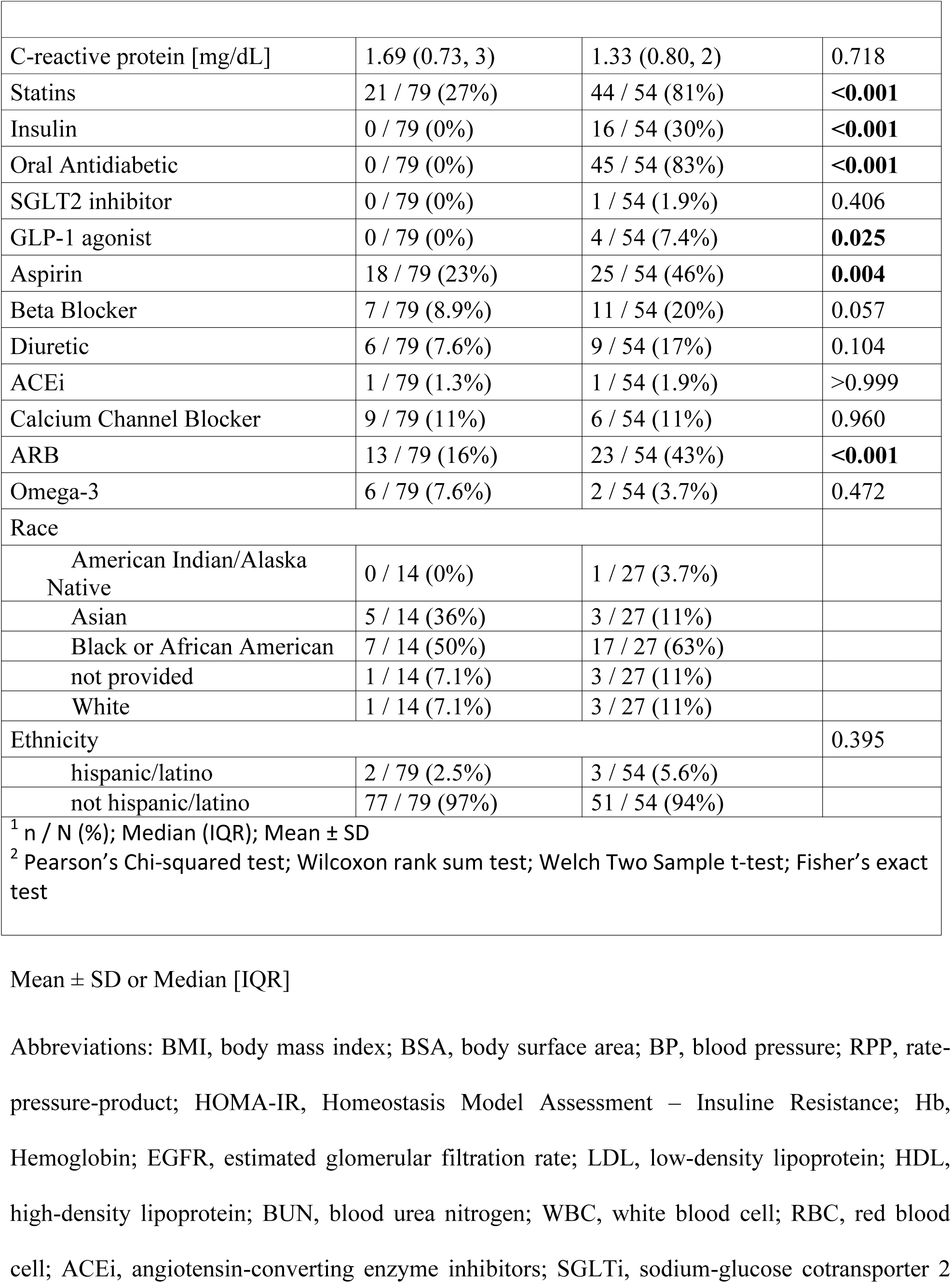
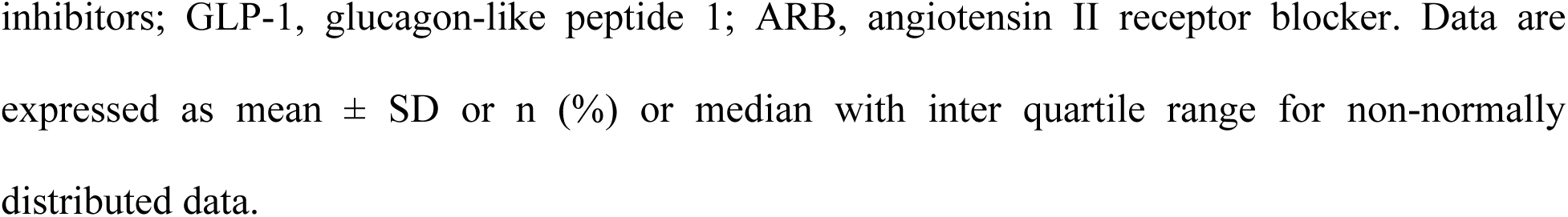
Participant Baseline Characteristics.

**Table 2.** Cardiac MRI Parameters, Vascular Reactivity and Cardiopulmonary Testing.

| Variable | Non T2DM | T2DM | p-value<sup>2</sup> |
| --- | --- | --- | --- |
| **CARDIAC MRI PARAMETERS** | **N = 55** | **N = 30** |  |
| LV EDVi [ml/m<sup>2</sup>] | 70 ± 15.0 | 69 ± 12.8 | 0.649 |
| LV ESVi [ml/m<sup>2</sup>] | 28 ± 8.2 | 28 ± 8.3 | 0.881 |
| LV ejection fraction [%] | 61 ± 5.7 | 60 ± 7.6 | 0.521 |
| LV global longitudinal strain [%] | -23 ± 4.9 | -22 ± 4.0 | 0.828 |
| LV mass index [g/m<sup>2</sup>] | 50 ± 13.6 | 50 ± 14.1 | 0.981 |
| LV mass/EDV ratio [g/ml] | 0.72 ± 0.19 | 0.75 ± 0.26 | 0.677 |
| Cardiac index [L/min/m<sup>2</sup>] | 2.79 ± 0.63 | 2.66 ± 0.57 | 0.355 |
| RV ESVi [ml/m<sup>2</sup>] | 30.46 ± 6.55 | 28.75 ± 8.65 | 0.351 |
| RV EDVi [ml/m<sup>2</sup>] | 66.97 ± 13.12 | 62.22 ± 16.18 | 0.173 |
| RV ejection fraction [%] | 54.38 ± 5.37 | 53.91 ± 5.94 | 0.719 |
| Rest MBF [ml/min/g] | 0.81 ± 0.18 | 0.80 ± 0.22 | 0.909 |
| Rest MBF (RPP-cor.) [ml/min/g] | 0.91 ± 0.23 | 0.88 ± 0.27 | 0.741 |
| Hyperemic MBF [ml/min/g] | 1.96 ± 0.53 | 1.77 ± 0.48 | 0.148 |
| Myocardial perfusion reserve | 2.47 ± 0.69 | 2.36 ± 0.72 | 0.581 |
| Myocardial native T1 [ms] | 1,120 ± 47 | 1,111 ± 38 | 0.423 |
| ECV | 0.28 ± 0.04 | 0.29 ± 0.03 | 0.420 |
| Any LV LGE | 1 (2.0%) | 0 (0%) | >0.999 |
| **VASCULAR REACTIVITY** | **N=67** | **N=40** |  |
| Resting brachial artery diameter (mm) | 4.2 ± 0.8 | 4.7 ± 0.8 | **0.003** |
| Flow Mediated Dilation (FMD)<br>(% of increase over baseline) | 6.5 ± 5.3 | 4.0 ± 5.0 | **0.018** |
| Nitroglycerin Induced Dilation (NID) (% of<br>increase over baseline) | 9.8 ± 4.8 | 5.9 ± 7.4 | **0.040** |
| Acetylcholine induced skin vasodilation (% of increase over baseline) | 50 ± 65 | 41 ± 84 | 0.531 |
| Sodium nitroprusside skin induced vasodilation (% of increase over baseline) | 77 ± 71 | 73 ± 78 | 0.772 |
| **CARDIOPULMONARY TESTING** | N = 39 | N = 20 |  |
| Peak VO<sub>2</sub> [ml/min/kg] | 20.7 ± 5.0 | 17.5 ± 4.8 | **0.022** |
| Peak VO<sub>2</sub>/WR [ml/min/W] | 9.8 ± 1.8 | 8.7 ± 1.7 | **0.017** |
| Peak respiratory exchange ratio (RER) | 1.11 ± 0.12 | 1.12 ± 0.10 | 0.741 |
| O<sub>2</sub> pulse peak (units?) | 12.1 ± 3.8 | 10.5 ± 3.1 | 0.098 |
| VE-VCO<sub>2</sub> slope (units ?) | 27.8 ± 3.6 | 28.2 ± 3.3 | 0.643 |
| Chronotropic index | 0.7 ± 0.3 | 0.8 ± 0.2 | 0.451 |

Mean ± SD
VO<sub>2</sub>/WR = Oxygen consumption (ml/min)/Work rate (watts) shows the relationship of oxygen consumption to work performed; RER = Respiratory Exchange Ratio, the volume of carbon dioxide produced to oxygen consumed (also known as RQ, respiratory quotient).
VE/VCO<sub>2</sub> slope = minute ventilation to carbon dioxide produced, a prognostic marker for either cardiac, pulmonary or both as limiting factors such as when there is ventilatory perfusion mismatch often associated with increased cardiac wedge pressure, pulmonary hypertension or pulmonary causes.
Abbreviations: LV, left ventricular; EDVi, enddiastolic volume index; ESVi, endsystolic volume index; RV, right ventricular; MBF, myocardial blood flow; ECV, extracellular volume.

### Cardiac MRI (CMR) Results

Due to COVID-19 restrictions, CMR was performed in a subset of 85 subjects (30 T2DM subjects). There were no differences in age or sex between subjects that underwent CMR and those who did not. We did not observe significant differences for any CMR measurements related to T2DM status, including rest (rMBF) and hyperemic myocardial blood flow (hMBF) (**Table 2).** Rest MBF was overall higher in females compared to males (p<0.05), also when normalized by the rate pressure product (RPP) (**Figure 1A)**. Hyperemic MBF was 0.39 ml/min/g lower in males with T2DM (p=0.05) compared to males without T2DM, and within the T2DM group it trended lower in males compared to females (p=0.089, **Figure 1B**). Hyperemic MBF correlated negatively with the log-transformed WBC (r=-0.29; p=0.013; **Figure 1C**). Interleukin-13, an anti-inflammatory type-2 cytokine with a putative protective role against atherosclerosis, correlated positively with hMBF (r=0.31; p=0.014, **Figure 1D**). Hyperemic MBF correlated negatively with age (r= –0.28; p=0.016). The extracellular volume (ECV) was higher in females (p=0.008), and T2DM did not have a significant effect (p=0.48). The LV ejection fraction at rest correlated negatively with ECV in females (r=-0.43; p=0.011), but not in males (r=-0.43; p=0.83; see **supplementary Figure 1**). Only one participant without T2DM had evidence of myocardial scar by LGE in the LV.

**Figure 1.**
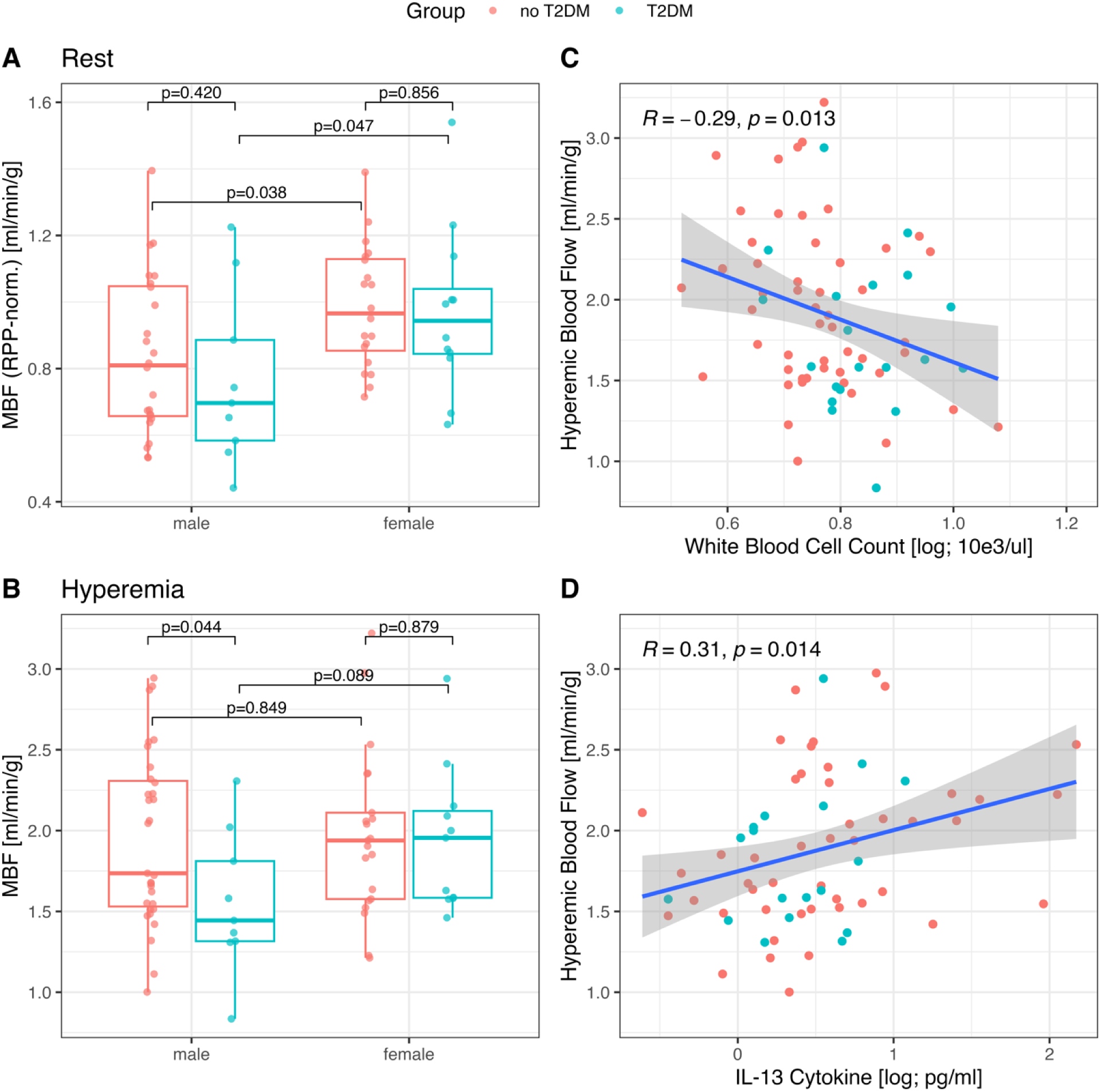
Changes in myocardial blood flow by T2DM status and markers of inflammation. **A**) Rest myocardial blood flow (MBF) was overall higher in female subjects compared to males (p=0.049), and this also applied to rest MBF normalized by rate pressure product. The presence of type-2 diabetes mellitus (T2DM) did not affect resting blood flow (p=0.318). **B)** MBF during hyperemia induced by regadenoson was lower in participants in males with T2DM (p=0.05), but not in females (p=0.179). The brackets show p-values from post-hoc pairwise comparisons of the marginal means between males and females within each patient group (with and without T2DM). **C)** The white blood cell count (WBC) was significantly higher in individuals with type-2 diabetes mellitus (5.50 (4.90, 7) vs. 6.50 (5.40, 8) pg/mL; p = 0.004) and correlated negatively with myocardial blood flow during hyperemia (r=-0.29; p=0.013). **D)** Interleukin-13, an anti-inflammatory type-2 cytokine with a putative protective role against atherosclerosis was not significantly different between participants with and without T2DM (p=0.34) but correlated positively with hyperemic (“stress”) MBF (r=0.33; p=0.008).

### CPET Results

CPET was performed in a cohort subset of 59 subjects (20 T2DM subjects). There were no differences in age or sex between the subjects that underwent CPET and those who did not. The respiratory exchange ratio (RER) averaged 1.12 ± 0.11, correlated with BMI (r=-0.49; p<0.001), and both T2DM and sex had no significant effect on RER. Peak VO2 was lower with T2DM (17.5±4.8 with T2DM vs. 20.7±5.0; p=0.022), mostly because of the effect of T2DM in females (14.2±2.9 with T2DM vs. 19.7±4.1 ml/min/kg; p<0.001, **Figure 2a**). The chronotropic index (CI) reached during CPET was similar in individuals with and without T2DM. The correlation of peak VO2 with the chronotropic index was noticeably different in males (r=0.56; p=0.001) compared to females (r=0.21; p=0.31; **Figure 2B**). The estimates of for the regression coefficients in the multivariable model for peak VO2 are shown in **supplemental Table 3**A. The model accounted for > 50% of the observed variability of peak VO2 and predicts that peak VO2 was nominally reduced by T2DM (p=0.096), lower in females (p<0.001), decreased with advancing age (p=0.003), and negatively associated with BMI in females (p=0.034) but not males. The effect of CI on peak VO2 was significantly different between males and females (p=0.007), consistent with **Figure 2B**. Peak O2 pulse, an index of stroke volume, was significantly lower in females compared to males, and T2DM had no significant effect. Peak O2 pulse correlated strongly with the stroke volume measured at rest with CMR (r=0.69; p<0.001), and with the myocardial ECV (r=-0.36; p=0.015), a marker of diffuse myocardial fibrosis. The association of ECV with peak O2 pulse was also significant (p=0.041) in a multilinear regression model for peak O2 including sex as shown in **supplementary Table 3B**.

**Figure 2.**
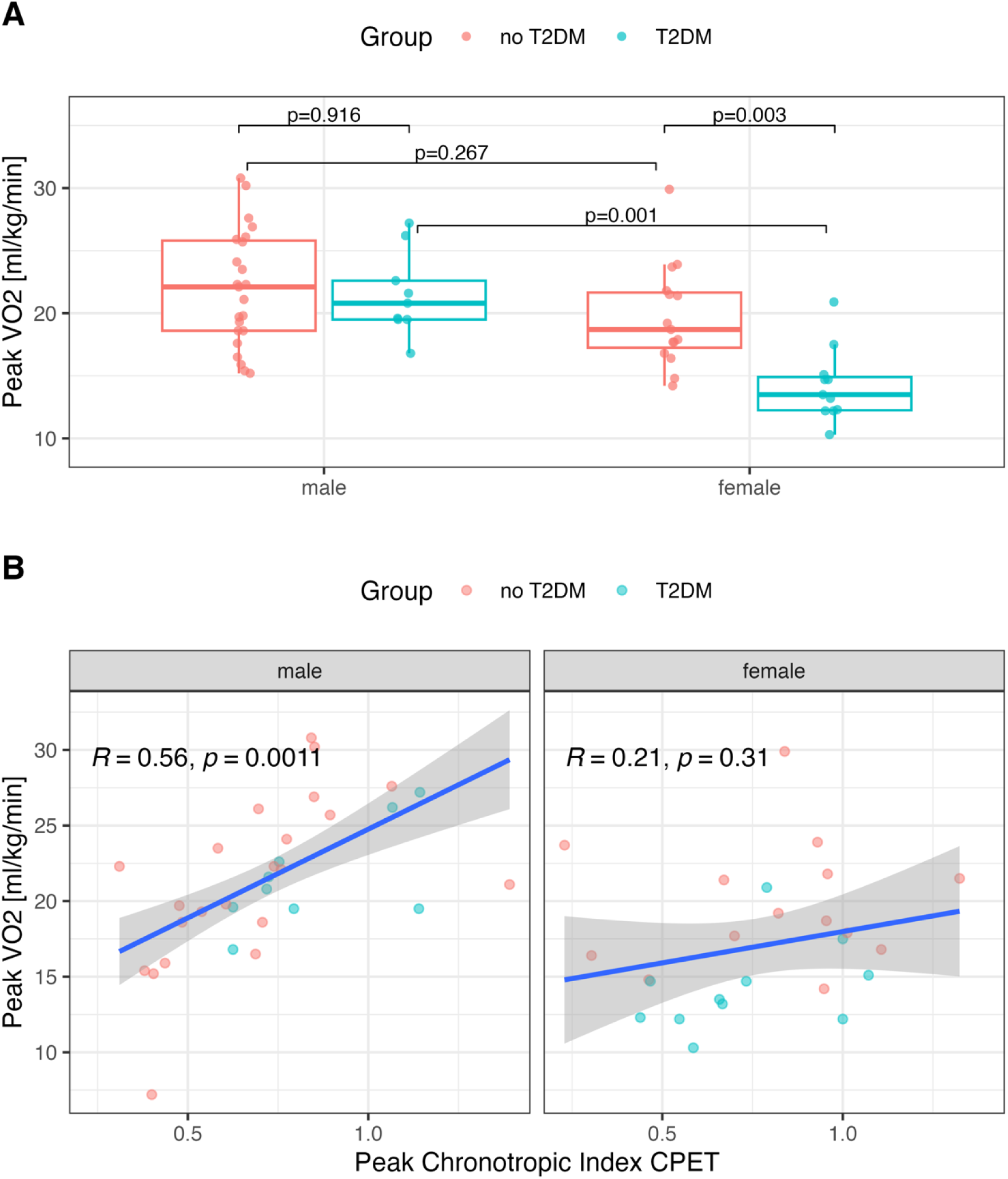
Cardiopulmonary Exercise Testing. **A**) Peak VO2 was reduced in T2DM females but there was no difference in males. **B)** Peak VO2 correlated significantly in males with the chronotropic index during CPET (r=0.056; p=0.001), but not in female participants (r=0.21; p=0.31). The chronotropic index (CI) is defined as the difference of peak heart rate during CPET and the resting heart rate, normalized by the age-adjusted expected heart response, defined as 220 – age [years] – resting heart rate.

The increase of the serum inflammatory markers IL-6 and IL-8 during CPET (value right after CPET – baseline value) was larger in participants with T2DM than without T2DM.

### Vascular Reactivity Results

FMD, a measure of vascular endothelial function, was lower in T2DM individuals compared to controls (Table 2). There was a significant interaction between T2DM and sex in their effects on FMD – FMD was reduced in males with T2DM (–4.44±1.36; p=0.002) compared to male controls, but the effect of T2DM was absent in females (p=0.972; **supplementary Figure 2A**). Similarly, nitroglycerin induced dilation (NID), a measure of vascular smooth muscle function, was reduced by –6.8±1.99 (p=0.001) in males with T2DM compared to males controls, but there was no effect of T2DM in females (p=0.768; **supplementary Figure 2B**). No differences were observed in the forearm skin vasodilation (endothelium-dependent and independent) between T2DM and controls. NID correlated with METs for vigorous plus moderate exercise (r=0.362, p=0.018) and METs for all activities (r=0.281, p=0.079). NID correlated nominally with hMBF (r=0.353; p=0.055). The correlation of FMD with hMBF did not reach statistical significance (r=0.23; p=0.083). Peak VO2 did not correlate significantly with FMD (r=0.20; p=0.178) and NID (r=-0.31; p=0.2).

### Serum and Skin Inflammatory Markers

White blood cell count (WBC) was increased in individuals with T2DM (p=0.01), mostly because WBC was higher in females with T2DM compared to female controls (p=0.001), while in males there was no significant effect of T2DM (p=0.474). A 2-way ANOVA model for the WBC confirmed this interaction between T2DM and sex (p=0.039), in addition to the main effect of T2DM (p=0.003). IL-5 was lower in T2DM participants (p=0.047 from Wilcoxon rank sum test; **Supplemental Table 2**). GDF15 was higher in T2DM individuals (p=0.012), and 2-way ANOVA model for GDF15 confirmed the effect of T2DM (p=0.008), while sex had no significant effect (p=0.21). No other major differences were observed between the groups among the inflammatory cytokines, adhesion molecules and growth factors in **Supplemental Table 2**. No differences were observed in the forearm skin biopsies in factors that included mast cell degranulation, number of vessels, macrophages, and round inflammatory cells (data not shown).

Peak VO2 correlated moderately with WBC (r= –0.540, p<0.0001), IL-4 (r=0.289, p=0.034), IL-5 (r=0.273, p=0.046) and IL-13 (r=0.418, p=0.002). VO2/WR correlated with ITAC (r= –0.317, p=0.023), IL-2, (r=0.282, p=0.045) and IL-13 (r=0.285, p=0.042), and there was weak evidence for a correlation with IL-4 (r=0.253, p=0.073).

## DISCUSSION

By employing state-of-the-art, detailed inflammatory phenotyping, noninvasive cardiac imaging and cardiopulmonary exercise testing, this observational study of elderly (>65 years) individuals explored the independent contributions of T2DM, age, sex and inflammation on cardiovascular health. Our findings in elderly persons with well-managed glycemic control and treatment of comorbidities indicate that older age, the combination of male sex and T2DM, and markers of systemic inflammation are factors contributing to impaired coronary and peripheral (micro)vascular function. Of note is the interaction of the effects of sex and T2DM on microvascular and cardiopulmonary function: both coronary and peripheral vascular function were reduced in males with T2DM and appeared to be largely preserved in females with T2DM, compared to elderly controls without T2DM. In contrast, aerobic exercise capacity was more impaired in females, compared to males, independent of T2DM status. Females had higher levels of adverse myocardial remodeling that may result in exercise-induced diastolic dysfunction.

Female sex was associated in previous studies with higher hMBF ^26,27^. The effect of DM on hMBF was not found to be significant in a sub-cohort aged 45 to 85 from the Multi-Ethnic Study of Atherosclerosis, and the authors did not report on any possible interaction of DM with sex^27^. In a study of premenopausal females with DM, hMBF and coronary perfusion reserve were lower in females with DM than without DM but still higher compared to nondiabetic older (∼15 years) post-menopausal females ^28^. This latter finding suggests that older age and change in menopause status can have effects on hMBF of similar magnitude as DM. In a PET study by Haas et al. of individuals with T2DM and good cardiometabolic control, both rest and hyperemic MBF were higher in females (aged 52±8 years) compared to males of similar age, yet the perfusion reserve was lower in females (menopausal status was not specified). In contrast, our study shows a similar myocardial perfusion reserve in elderly males and (postmenopausal) females without T2DM, but hMBF was significantly lower in males with T2DM. Our results suggest that menopause does not blunt the differences of hMBF and coronary reactivity seen in pre-menopausal females, compared to males. Any benefits of better vascular function after menopause, may also lessen the impact of T2DM on coronary vascular function and result in a more pronounced effect on T2DM on coronary microvascular function in males compared to females.

An impaired MBF may be confounded by the presence of obstructive coronary lesions ^14,29^. Only one participant without T2DM showed LGE in the LV, which suggests that obstructive coronary lesions were not a major factor. The consistency of a male-specific effect of T2DM on hMBF, and brachial artery FMD and NID, and the low probability of simultaneous flow-limiting stenosis in both vascular beds without symptoms suggest that obstructive disease was not a major confounder.

The measures of peripheral endothelial and smooth muscle vascular function obtained in our study (FMD, NID) provide additional evidence for similar male-specific effects of T2DM as for the coronary hyperemic response. In a study assessing reference intervals and risk factors for FMD across a broad age-range, Holder et al found some evidence of an interaction of sex with DM: age– and sex-specific Z scores for FMD were lower in unmedicated males with DM, but not females (p=0.08 for interaction effect), while in individuals taking medications DM had an effect in both sexes with no evidence of an interaction of sex with DM ^30^. Since our study included only individuals aged ≥65 years it provides evidence of an interaction of sex with T2DM without potential confounding by age. Holder et al, who enrolled predominantly individuals aged <65 years, reported significantly higher FMD in females compared to males up to the age of 70 years, while in older persons this difference disappeared. This latter finding is consistent with our observation that FMD was not different between elderly males and females *without* T2DM.

We found no major differences between T2DM and controls in serum and skin inflammatory markers, apart from increased WBC and lower IL-5 levels among those with T2DM. GDF15 was higher among those with T2DM however this is likely explained by metformin increasing GDF15 levels ^31^. WBC correlated with coronary blood flow, which is in accordance with previous studies that have shown that individuals with elevated WBC have reduced epicardial coronary flow and are at higher risk of acute myocardial infarction ^32^. A high WBC also mediates insulin resistance and predicts the development of T2DM ^33^.

Previous studies in individuals with both type 1 and 2 diabetes mellitus have shown that cardiac autonomic neuropathy leading to inability to increase heart rate during exercise was associated with a reduced peak VO2 ^34^. During exercise, the healthy heart increases both stroke volume and heart rate. When the cardiac contractility reserve is impaired (e.g. with incipient heart failure), peak VO2 fails to increase with heart rate as the stroke volume becomes the limiting factor. In our study, peak VO2 did not increase significantly with the CI in females (**Figure 2B**) suggesting that any chronotropic incompetence would be less of a limiting factor than in men. The sex-related difference in the relation of peak VO2 with CI was confirmed in a multivariable analysis (**supplemental Table 3A**). We also noted a negative correlation in females of ECV with the resting LV ejection fraction and peak O2 pulse (**supplemental Table 3B**). Diffuse myocardial fibrosis can impair augmentation of relaxation velocity as heart rate increases during exercise ^35^. This suggests that diffuse myocardial fibrosis may cause diastolic dysfunction with exertion not detectable at rest. The stroke volume fails to rise with heart rate, and patients experience dyspnea and fatigue. As ECV was significantly higher in female participants compared to males, effects of diffuse fibrosis on cardiac function may manifest more readily in elderly females. The prevalence of LV diastolic dysfunction markedly increases after menopause and may lead to heart failure ^36^. The moderately strong negative correlation between peak VO2 and WBC measured at rest suggests a link between reduced exercise capacity and systemic inflammation. Nevertheless, T2DM may exacerbate the effects of inflammation as acute maximal exercise effort caused a larger transient increase in IL-6 and IL-8 in T2DM compared to controls.

Our study has several limitations. Owing to the constraints imposed by the COVID-19 pandemic, not all participants could complete the entire range of planned examinations. The number of participants with T2DM was lower than the number of without T2DM, however the number of individuals studied was sufficient to provide adequate statistical power for group comparisons. Our study population had a mean duration of diabetes of 15 years and a mean HbA1c of 7 % which reflects good glycemic control for an elderly population ^37^. Our subjects with T2DM had very prevalent use of statins and anti-hypertensive medications, indicating that cardiovascular comorbidities were well controlled. With respect to diabetes management, at the time we conducted our study, glucagon-like peptide 1 (GLP1) receptor agonists and SGLT2 inhibitors were not used as commonly as they are now. Given their cardiovascular benefits in T2DM ^38,39^, our results might underestimate the cardiovascular health of T2DM participants currently using these drugs. Our findings may not be generalizable to those with diabetes of much longer or shorter duration, worse glycemic control, or suboptimal management of cardiovascular co-morbidities. Last, our data on physical activity was self-reported. Although we used a validated physical activity assessment ^40^, there could be bias in self-reported physical activity data by sex and/or by T2DM/control status.

### Conclusions

In elderly individuals, whose CVD risk factors are managed according to contemporaneous guidelines, T2DM has a sex-specific effect on coronary and peripheral vascular phenotypes, impairing vascular function and reactivity predominantly in men. In contrast, aerobic exercise capacity was more impaired in females, compared to males, independent of T2DM status, and females showed higher levels of adverse myocardial remodeling that may result in exercise-induced myocardial dysfunction. Our findings suggest that the pathophysiological manifestations of T2DM on vascular function and exercise capacity are distinct in elderly males and females and this may reflect underlying differences in vascular and myocardial aging in the presence of T2DM.

## Data Availability

All data are available

## SUPPLEMENTAL DATA

**Supplemental Table 1:** CMR Measurements Stratified by Sex.

| Variable | Females |  |  | Males |  |  |
| --- | --- | --- | --- | --- | --- | --- |
|  | no T2DM<br>N = 22 | T2DM<br>N = 18 | p-value | no T2DM<br>N = 33 | T2DM<br>N = 12<sup>1</sup> | p-value |
| LV EDVi [ml/m<sup>2</sup>] | 68 ± 12.6 | 66 ± 12.4 | 0.517 | 72 ± 16.5 | 74 ± 12.5 | 0.672 |
| LV ESVi [ml/m<sup>2</sup>] | 27 ± 8.0 | 26 ± 8.3 | 0.524 | 28 ± 8.4 | 32 ± 7.4 | 0.180 |
| LV ejection fraction [%] | 60 ± 6.4 | 61 ± 8.0 | 0.604 | 61 ± 5.3 | 57 ± 6.2 | 0.063 |
| LV GLS [%] | -22 ± 4.9 | -23 ± 4.0 | 0.370 | -23 ± 5.0 | -21 ± 3.7 | 0.176 |
| LV mass index [g/m<sup>2</sup>] | 44 ± 11.9 | 44 ± 7.7 | 0.918 | 54 ± 13.3 | 61 ± 16.4 | 0.235 |
| LV mass/EDV ratio [g/ml] | 0.64 ± 0.12 | 0.70 ± 0.16 | 0.260 | 0.78 ± 0.21 | 0.82 ± 0.36 | 0.736 |
| Cardiac index [L/min/m<sup>2</sup>] | 2.68 ± 0.69 | 2.63 ± 0.64 | 0.818 | 2.87 ± 0.57 | 2.72 ± 0.46 | 0.378 |
| Rest MBF [ml/min/g] | 0.85 ± 0.17 | 0.88 ± 0.21 | 0.652 | 0.78 ± 0.18 | 0.69 ± 0.19 | 0.218 |
| Rest MBF (RPP-cor.) [ml/min/g] | 0.99 ± 0.18 | 0.97 ± 0.25 | 0.856 | 0.84 ± 0.24 | 0.77 ± 0.26 | 0.482 |
| Hyperemic MBF [ml/min/g] | 1.97 ± 0.51 | 1.94 ± 0.45 | 0.872 | 1.95 ± 0.54 | 1.55 ± 0.44 | **0.039** |
| Myocardial native T1 [ms] | 1,133 ± 51 | 1,118 ± 39 | 0.350 | 1,108 ± 41 | 1,103 ± 37 | 0.698 |
| ECV | 0.30 ± 0.03 | 0.29 ± 0.03 | 0.675 | 0.27 ± 0.03 | 0.29 ± 0.05 | 0.306 |
| Any LV LGE | 0 (0%) | 0 (0%) | >0.999 | 1 (3.3%) | 0 (0%) | >0.999 |

Mean ± SD
Abbreviations: LV: left ventricular; EDVi: enddiastolic volume index; ESVi: endsystolic volume index; GLS: global longitudinal strain; RV: right ventricular; MBF: myocardial blood flow; ECV: extracellular volume; LGE: late gadolinium enhancement.

**Supplemental Table 2:** Biomarkers of inflammation and oxidative stress.

| Variable | no T2DM, N = 79 | T2DM, N = 54 | P-value |
| --- | --- | --- | --- |
| C-reactive protein [mg/l] | 1.69 [0.73, 2.84] | 1.33 [0.80, 2.44] | 0.718 |
| WBC [10e6/ul] | 5.50 [4.90, 6.65] | 6.50 [5.40, 7.60] | **0.004** |
| insulin_uIU_ml | 6 [4, 9] | 10 [6, 20] | **<0.001** |
| ITAC_pg/ml | 12 [8, 18] | 12 [9, 21] | 0.385 |
| GM-CSF_pg/ml | 3.9 [1.7, 7.5] | 3.6 [2.1, 6.6] | 0.959 |
| Fractalkine_pg/ml | 58 [25, 78] | 56 [40, 72] | 0.971 |
| INF-g_pg/ml | 9 [3, 13] | 8 [5, 13] | 0.886 |
| IL-10_pg/ml | 3.6 [2.2, 7.9] | 3.9 [2.0, 7.4] | 0.570 |
| MIP-3a_pg/ml | 4.99 [3.29, 7.03] | 5.51 [3.82, 7.55] | 0.338 |
| IL-12 (p70)_pg/ml | 1.48 [0.93, 2.49] | 1.32 [0.88, 2.31] | 0.506 |
| IL-13_pg/ml | 3 [1, 7] | 2 [1, 5] | 0.405 |
| IL-17A_pg/ml | 3.39 [2.05, 5.52] | 2.89 [2.05, 4.71] | 0.690 |
| IL-1b_pg/ml | 0.52 [0.41, 0.87] | 0.52 [0.38, 0.75] | 0.334 |
| IL-2_pg/ml | 1.57 [0.75, 2.34] | 1.36 [0.93, 1.86] | 0.471 |
| IL-21_pg/ml | 2.42 [1.12, 4.01] | 2.83 [1.56, 4.20] | 0.659 |
| IL-4_pg/ml | 20.6 [6.8, 37.8] | 12.9 [7.0, 34.5] | 0.517 |
| IL-23_pg/ml | 57.4 [31.4, 106.9] | 39.1 [28.5, 73.0] | 0.187 |
| IL-5_pg/ml | 1.55 [0.59, 2.67] | 0.83 [0.44, 2.03] | **0.047** |
| IL-6_pg/ml | 2.9 [1.2, 7.9] | 1.9 [0.8, 5.1] | 0.095 |
| IL-7_pg/ml | 16.3 [5.9, 22.8] | 15.5 [12.1, 19.0] | 0.519 |
| IL-8_pg/ml | 5.6 [3.0, 8.1] | 6.6 [2.8, 13.8] | 0.417 |
| MIP-1a_pg/ml | 7.9 [3.4, 9.6] | 7.7 [6.2, 9.4] | 0.666 |
| MIP-1b_pg/ml | 7.5 [5.2, 11.0] | 9.0 [6.4, 12.6] | 0.153 |
| TNF-a_pg/ml | 3.94 [2.04, 4.79] | 4.24 [3.41, 5.32] | 0.122 |
| EGF_pg/ml | 14 [7, 31] | 8 [5, 26] | 0.174 |
| FGF-2_pg/ml | 42 [11, 115] | 59 [11, 109] | 0.951 |
| VEGF_pg/ml | 55 [1, 319] | 113 [1, 523] | 0.460 |
| MMP-2 [ng/ml] | 138 [127, 155] | 135 [125, 143] | 0.182 |

| **Variable** | **no T2DM, N = 79** | **T2DM, N = 54** | **P-value** |
| --- | --- | --- | --- |
| MMP-9 [ng/ml] | 78 [49, 121] | 75 [66, 145] | 0.365 |
| ADAMTS13_pg/ml | 2,316 [1,641, 3,578] | 2,619 [1,695, 4,116] | 0.242 |
| D-Dimer [ng/ml] | 3.0 [1.3, 8.2] | 3.5 [1.5, 7.7] | 0.930 |
| GDF15_pg/ml | 2.1 [1.2, 5.2] | 3.9 [1.9, 5.7] | **0.012** |
| Myoglobin_pg/ml | 159 [83, 245] | 145 [71, 257] | 0.477 |
| sICAM-1_pg/ml | 183 [121, 844] | 207 [124, 364] | 0.782 |
| MPO_pg/ml | 251 [142, 556] | 306 [153, 567] | 0.502 |
| P-Selectin_pg/ml | 194 [131, 328] | 179 [125, 308] | 0.879 |
| NGAL_pg/ml | 152 [91, 281] | 161 [102, 320] | 0.521 |
| sVCAM-1_pg/ml | 1,428 [877, 2,857] | 1,415 [886, 2,729] | 0.867 |
| SAA [ng/ml] | 17 [9, 49] | 21 [14, 193] | 0.194 |

Median [IQR]
Abbreviations: WBC, white blood cell count; ITAC, Interferon–inducible T Cell Alpha
Chemoattractant; GM-CSF, Granulocyte macrophage colony-stimulating factor; INF-g,
Interferon gamma; IL, Interleukin; MIP, Macrophage Inflammatory Protein; TNF-a, tumor
necrose factor alpha; EGF, epidermal growth factor; FGF, fibroblast growth factor; VEGF,
vascular endothelial growth factor; MMP, matrix metalloproteinase; ADAMTS, von Willebrand
factor-cleaving protease; GDF, Growth/Differentiation Factor; sICAM, soluble intracellular
adhesion molecule; MPO, myeloperoxidase; NGAL, Neutrophil Gelatinase-Associated
Lipocalin; sVCAM, soluble Vascular Cell Adhesin Molecule; SAA, Serum amyloid A protein.

**Supplemental Table 3A:** Coefficient estimates from linear multi-variable regression model for peak VO2. Continuous predictors were centered and scaled by their standard deviation. **3B**: Coefficient estimates from linear multi-variable regression model for peak O2 pulse. The number of observations used for this model (N=42) was smaller than for peak VO2 because it required paired measurements from CMR and CPET.

**A) Linear Model for CPET Peak VO<sub>2</sub>**
| *Predictors* | **Peak VO<sub>2</sub> [ml/kg/min]** |  |  |
| --- | --- | --- | --- |
|  | *Estimates* | *CI* | *p* |
| (Intercept) | 22.10 | 20.78 – 23.43 | **<0.001** |
| Chronotropic Index [1 SD] | 3.44 | 2.12 – 4.76 | **<0.001** |
| Female sex | -5.35 | -7.43 – -3.28 | **<0.001** |
| T2DM | -1.77 | -3.87 – 0.33 | 0.096 |
| Age [Y] | -2.04 | -2.98 – -1.11 | **<0.001** |
| BMI [1SD] | -0.69 | -2.39 – 1.02 | 0.422 |
| Female sex : Chronotropic Index | -2.65 | -4.52 – -0.77 | **0.007** |
| Female sex : BMI | -2.57 | -4.95 – -0.20 | **0.034** |
| Observations | 55 |  |  |
| R<sup>2</sup> / R<sup>2</sup> adjusted | 0.666 / 0.616 |  |  |

**B) Linear Model for Peak O2 Pulse**
| *Predictors* | **Peak O2 Pulse [ml/beat]** |  |  |
| --- | --- | --- | --- |
|  | *Estimates* | *CI* | *p* |
| (Intercept) | 14.66 | 13.63 – 15.70 | **<0.001** |
| T2DM | -1.27 | -3.15 – 0.60 | 0.176 |
| Female sex | -4.54 | -6.34 – -2.75 | **<0.001** |
| BMI [1 SD] | 1.17 | 0.24 – 2.09 | **0.015** |
| Age [1 SD] | -0.44 | -1.09 – 0.22 | 0.186 |
| Myocardial ECV [1 SD] | -0.87 | -1.56 – -0.17 | **0.016** |
| Chronotropic Index [1 SD] | -0.81 | -1.44 – -0.17 | **0.014** |
| Female Sex : T2DM | -0.36 | -2.97 – 2.26 | 0.783 |
| Observations | 42 |  |  |
| R<sup>2</sup> / R<sup>2</sup> adjusted | 0.818 / 0.781 |  |  |

**Supplemental Figure 1:**
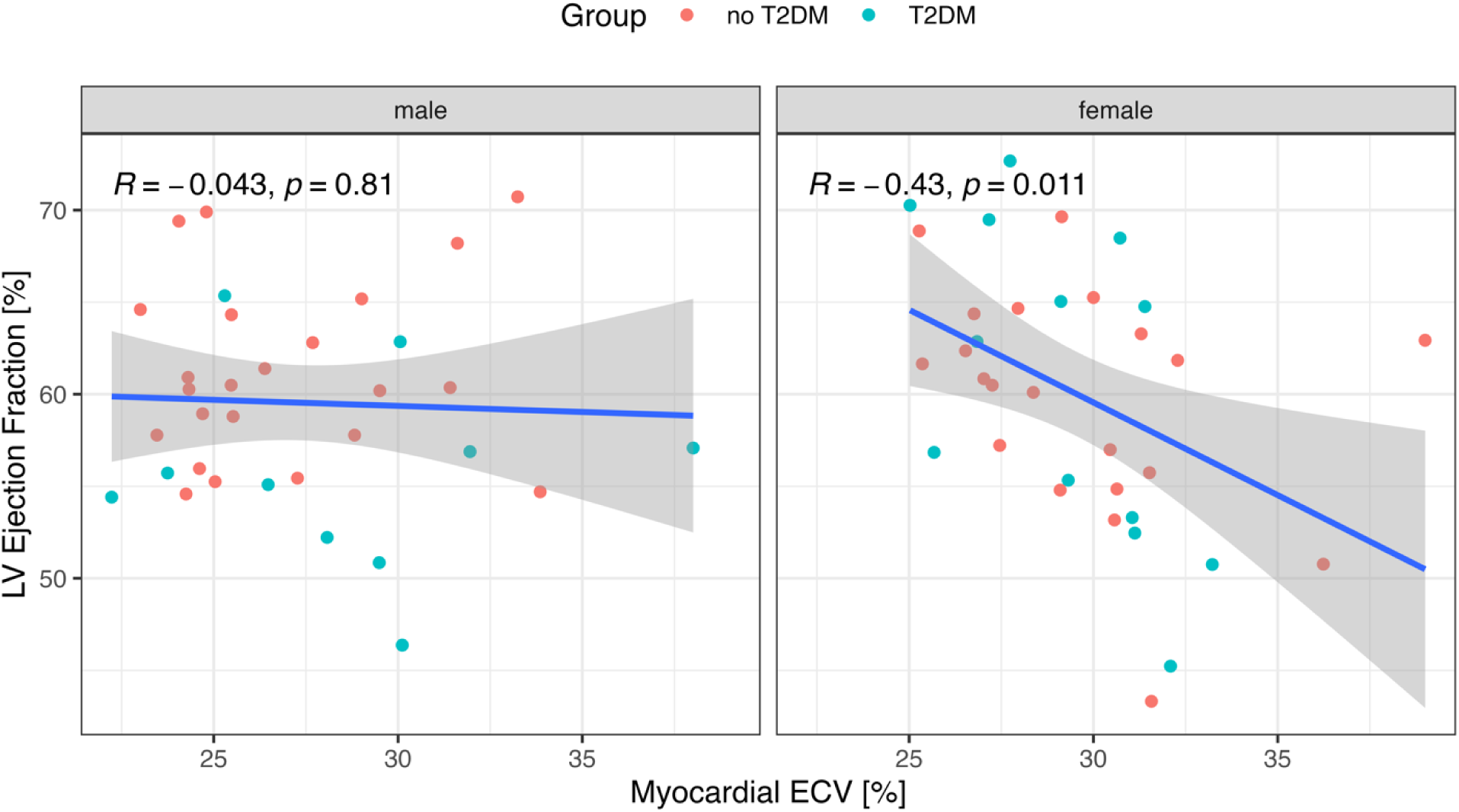
Myocardial extracellular volume (ECV), a biomarker of diffuse fibrosis when infiltrative disease can be excluded, correlated significantly in females with the LV ejection fraction at rest, but not in males. Both ECV and LV EF were quantified by cardiac magnetic resonance imaging.

**Supplemental Figure 2A:**
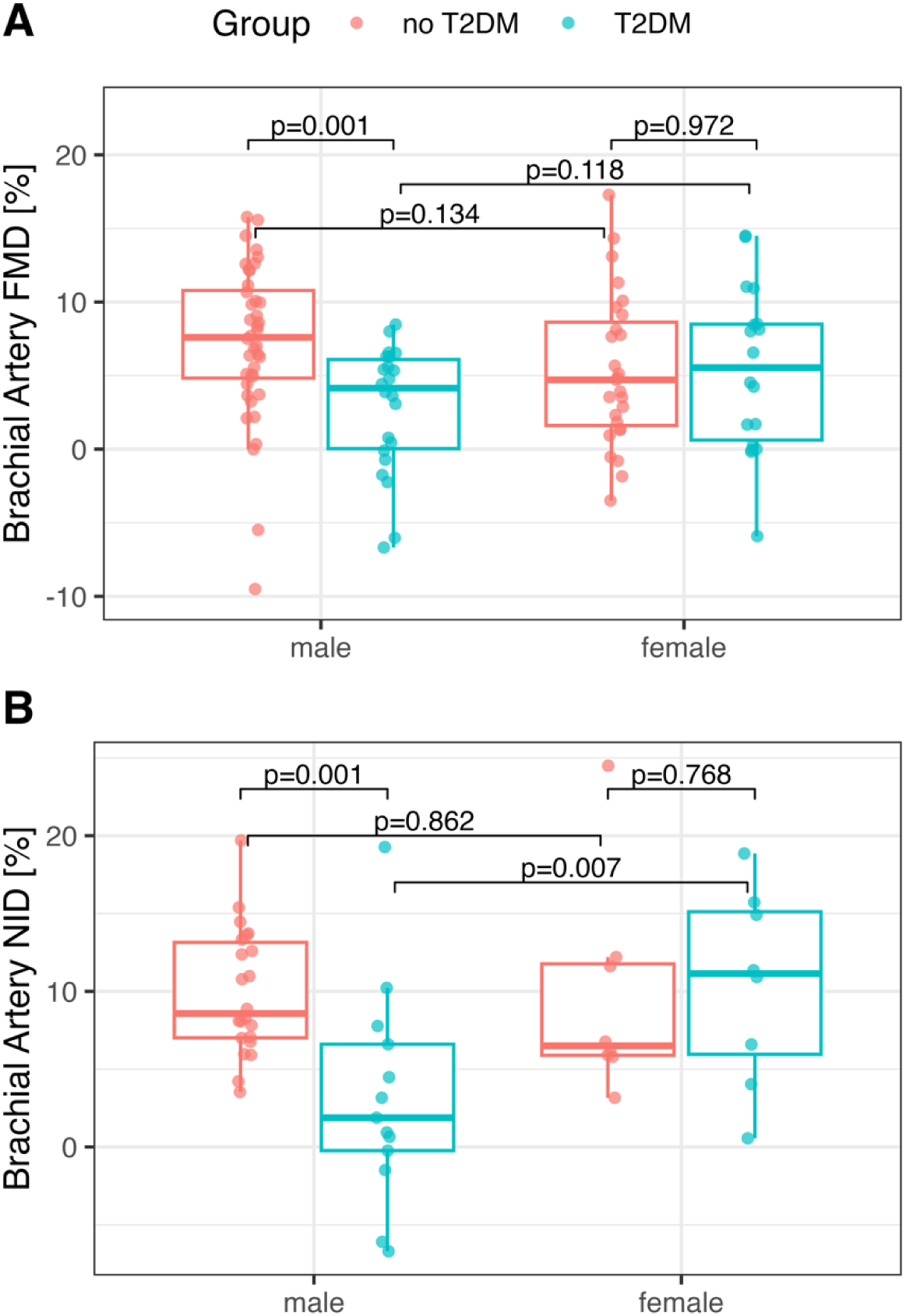
In an ANOVA model for the brachial artery flow mediated dilation (FMD), a measure of endothelial function, there was a significant interaction of sex with T2DM (p=0.032). Post-hoc pairwise comparisons with Tukey’s adjustment of p-values indicated that FMD was significantly lower in males with T2DM compared to males of similar age without T2DM (p=0.001), while in females there was no significant effect of T2DM (p=0.72). **2B**: A similar pattern of interaction of sex with T2DM was observed for nitroglycerin-induced vasodilation (NID), indicating that vascular smooth muscle function was abnormal in males with T2DM, compared to elderly males without T2DM (p=0.001), and also elderly females with T2DM (p=0.007).

